# Gene-environment interplay in the relationship between the visibility of the environment and self-reported depression in early midlife: a Finnish twin cohort study

**DOI:** 10.1101/2025.05.23.25328215

**Authors:** Zhiyang Wang, Stephanie Zellers, Maarit Piirtola, Sari Aaltonen, Jessica Salvatore, Danielle Dick, Simone Kühn, Jaakko Kaprio

## Abstract

**Background:** Depression is a major public health concern with a complex etiology, which may also be influenced by the living environment. The first-person visibility represents the most direct way of influence from the physical environment.

**Objectives:** We aim to investigate the effect of the visibility of the environment at the residence on self-reported depression among early midlife adults, as well as underlying gene-environment interplay.

**Methods:** The study used 1867 participants who completed the early midlife follow-u p (mean age: 37.2 years) of the FinnTwin12 cohort. The visibility of the environment at the residence was segmented into three visibility factors: sky, tree, and building. Linear regression was fitted between visibility factors and self-reported depression. Molecular methods using polygenetic risk score and twin design (univariate modeling and bivariate moderation modeling) were applied to explore the gene-environment correlation and interaction.

**Findings:** In males, a higher proportion of building visibility factor was associated with more depression (beta: 0.20, 95% CI: 0.03, 0.36). Univariate twin models estimated that additive genetic factors accounted for 46%, 29%, and 49% of the variance in sky, tree, and building visibility factors, respectively. Bivariate moderation analyses revealed strong gene-environment interactions between all three visibility factors and self-reported depression in females, while in males, interaction was observed by the building visibility factor.

**Conclusion:** Urban design should consider building density and other related characteristics to promote mental well-being. The complex gene-environment interplay informs the need for tailored interventions that account for genetic susceptibility and processes by which persons select their inhabitats.

## 1 Introduction

Depression is a serious public health burden, ranging from mild, temporary episodes of anhedonia and sadness to severe, persistent depression. According to the Global Burden of Disease Study 2019, the age-standardized prevalence of depressive disorder was 3.8%, being slightly higher in Western Europe (GBD 2019 Mental Disorders Collaborators, 2022). Depression is a common comorbidity of other psychological and psychiatric conditions such as anxiety or schizophrenia (Hartley et al., 2013) and is associated with unhealthy lifestyles and their consequences including smoking, alcohol dependence, and obesity (McCarty et al., 2009). These connections could be attributed to shared genetic factors, overlapping biological pathways, and so on(Gold et al., 2020). Twin and molecular studies showed that both genetic and environmental factors contribute to depression, highlighting its complex etiology (Adams et al., 2025; Polderman et al., 2015).

Visual perception is one of the most direct and effective ways for people to be influenced by the external environment (Hou et al., 2024a). Conventional quantification of the physical environment, i.e., satellite-based, cannot distinguish specific characteristics, such as whether green space consists of tree canopies, grass, or open areas, or whether tall buildings obstruct views, because it takes a birds-eye perspective (Kühn et al., 2023). For this reason, the analysis of what an individual can see from a first-person perspective might be crucial. The prospect-refuge theory suggests that open views offer a sense of safety and well-being (Appleton, 1975), which is beneficial to mental health. The stress reduction theory suggests that environmental exposures, such as green spaces, can alleviate the damage from external adverse stimulation through the reduction of the level of stress people experience (Ulrich et al., 1991). Highly occluded views hinder the visual connection to the sky and the surroundings, which impairs spatial orientation and induces stress (Chung et al., 2022a). Some types of views were associated with reduced noise annoyance, likely by means of restoration from stress (Chung et al., 2022b). The association between stress and depression is well-established (Tafet & Nemeroff, 2015). Moreover, the sky visibility factor and green view index were associated with thermal sensation and satisfaction(Liu et al., 2024), which are able to directly affect mental and emotional state (Yang et al., 2024). In a Dutch study of 95 children, grey matter volume in prefrontal clusters was associated with green open space coverage as well as sky view, implicating a neurological pathway (Kühn et al., 2023).

Genes also play a role in these mechanisms, and the relationship between genetic and environmental factors is complex, generally with two types: gene-environment correlation (rGE) and gene-environment interactions (G×E). rGE refers that heritable characteristics are correlated with the environment individuals face, with generally three subtypes: passive, where both genes and environment are provided by the parents; evocative, where genetically influenced traits evoke environmental specific responses; and active, where individuals actively seek out environments that align with their genetic predispositions (Dick, 2011). G×E describes that exposures may trigger, compensate, enhance, or socially control genetic influences on a phenotype (Shanahan & Hofer, 2005). Studying both types informs the causal mechanisms of psychopathology, identifies high-risk subgroups, optimizes current interventions, even in highly heritable diseases, and avoids misleading types of biological reductionism and stigma (Ordovas & Tai, 2008; Rutter et al., 2006). A few twin studies have already been conducted in this field with the outcome of depression. The twin design decomposes phenotypic variance into additive (A) and dominant (D) genetic components and common (C) and unique (E) environmental components. This approach estimates the genetic predisposition to exposures and allows the estimation of variables of interest varied by exposure to study gene-environment interplay. For example, in UK, significant genetic correlations between family environment (family chaos and parenting style) and depressive symptoms were observed among children, along with some evidence of G×E effects (Wilkinson et al., 2013). However, external physical exposures were not considered in such studies, since these exposures are usually regarded as non-genetic. We believe that people select their living environment, and factors influencing these choices may be shaped by genetic predispositions, making certain external physical exposures partially heritable.

Early midlife is a pivotal but relatively overlooked stage of life characterized by significant changes in health, cognition, and well-being, during which individuals take on key responsibilities of caregiving, supporting, and creating in the family and society (Lachman et al., 2014). In this study, we aim to elucidate the complex relationship between the visibility of the environment at the residence and self-reported depression among early midlife adults with three objectives: 1) assessing the environmental impact by visibility factors on self-reported symptoms of depression; 2) examining the influence of genetic predispositions on visibility factors; 3) investigating how the difference in visibility factors between cotwins moderates the genetic predisposition to self-reported symptoms of depression.

## 2 Method and materials

### 2.1 Participants

Participants were from the FinnTwin12 cohort, a nationwide prospective cohort of all Finnish twins born between 1983 and 1987. At baseline, 5,184 twins were enrolled when they reached age 11/12 years, and there were four survey follow-ups: age 14 years, age 17 years, young adulthood (mean age 24.2), and early midlife (mean age 37.2 years), with retention rates of 92%, 75%, 66%, and 41%, respectively. The early midlife follow-up was conducted in 2022 with a total of 2,122 participants. In addition to survey follow-ups, 1,035 families with 2,070 twins were invited to an intensive study involving psychiatric interviews, biological sampling, and additional questionnaires and tests at the follow-ups of age 14 years and young adulthood, yielding a total of 1,478 individual twins with genomic data. A recent study has described the latest follow-up of the FinnTwin12 cohort (Cooke et al., 2024).

### 2.2 Measures

#### 2.2.1 Self-reported depression

Self-reported depression was measured by the short-version Finnish-translated Center for Epidemiologic Studies Depression Scale (CES-D) (Saari et al., 2024), included in the early midlife follow-up questionnaire. This version of CES-D consists of eight items, each rated on a 4-point Likert scale ranging from 1 (rarely or none of the time) to 4 (most or all the time). Mean scores were calculated, allowing for up to two missing items. A higher score reflects more depressive symptoms. The Cronbach alpha for the scale reliability were 0.82.

#### 2.2.2 The visibility of the environment at the residence

Residential history, derived from the Digital and Population Data Services Agency, Finland, was used to to merge the visibility of the environment at the residence. It contains residential geocodes on the north and east coordinates and the age moving in and out of each address from birth to 2020. Concretely, the GSV2SVF tool was then used to segment the full-view panoramic photographs from Google Street View, as the first-person perspective, around participants’ residential address (200 m radius) (Liang et al., 2020). This tool uses the Caffe-SegNet deep convolutional framework to classify street photographs into three visibility factors: sky, trees, and buildings, and transforms them into fisheye images to represent the areal proportions of the fisheye image with a range between 0 and 1, as the visibility of the environment (Badrinarayanan et al., 2017). These factors denote the proportion of the hemisphere attributed to the sky, trees, and buildings when viewing from a ground location (Liang et al., 2020). Details on the procedure have been introduced in a previous publication (Kühn et al., 2023). Google Street View does not provide historical data; therefore, we had to take images at the point of analysis (median year: 2019, mean year: 2016). We merged visibility factors with participants’ residential addresses from 2019. If the 2019 address was unavailable, we substituted it with the 2018 address, and if still unavailable, with the 2020 address. This ensured that environmental exposure measurements were aligned with the period close to 2019, a few years before the early midlife follow-up. Since urbanization is relatively slow in Finland at present, this relieves some concerns regarding potential temporal mismatches (Evers et al., 2024).

#### 2.2.3 Polygenic risk score (PRS)

We used 12 PRS related to depression, including schizophrenia, body mass index, Alzheimer’s disease, neuroticism, broad depression, subjective well-being, major depressive disorder, insomnia, alcohol dependence, externalizing problems, smoking initiation, and educational attainment (Supplemental Table 1). PRSes were calculated using HapMap3 single-nucleotide polymorphisms with 1000 Genomes European minor allele frequency > 5%, using GWAS summary statistics for the effects of linkage disequilibrium (Privé et al., 2022). The reference panel consisted of 27,284 individuals from the FINRISK study (no overlap with the FinnTwin12 study). To generate a multi-PRS score, 12 PRSes were combined using the linear lasso penalized with self-reported CES-D score as the outcome variable (Albiñana et al., 2023). Sex, age at the early midlife follow-up, and principal components for ancestry were included as unpenalized terms in the model.

#### 2.2.4 Comparison variables

To compare the novel first-person perspective visibility factor and conventional satellite-based measures at the residence, two comparison variables, tree cover density and built-up areas (the extent of soil sealing where (semi)natural land cover is replaced by artificial surfaces), were selected. Both were obtained from the high-resolution Copernicus Land Monitoring Service (10 m) and were within a 100 m buffer around each geocode in 2018. We initially merged data using participants’ 2018 addresses. If the 2018 address was unavailable, we used the 2019 address, and if that was also unavailable, we used the 2020 address. The satellite data represents a bird-eye view of surroundings, rather than what individuals actually see when living in the respective neighborhood.

#### 2.2.5 Covariates

Several demographic and lifestyle variables were identified as covariates *a priori*: sex (categorical, female vs. male), age at the early midlife survey (continuous, year), work (categorical, not working or other situation vs. currently working), education (categorical, post-secondary or lower vs. bachelor/equivalent or above), living status (categorical, a spouse/partner vs. a spouse/partner and child(ren) vs. alone vs. other), illicit substance ever use (categorical, never vs. at least once), ever smoker (smoked over 100 cigarettes lifetime; categorical, never vs. ever smoker), alcohol drinking (categorical, at most monthly vs. 2-4 times a month vs. 2-3 times a week or more), and leisure time physical activity (categorical, once a week or less vs. 2–3 times a week vs. 4–5 times a week or more). Sex was assigned at birth, and other variables were inquired in the early midlife questionnaire. Additionally, to account for the social environment, three neighborhood social variables at the postal code level were derived from Statistics Finland (www.stat.fi) in 2019: the proportion of residents with the lowest education level, of residents with the lowest income quartile, and of unemployed residents (Kivimäki et al., 2018). The merging rule was the same as for visibility factors. A neighborhood deprivation score was calculated by averaging the z-scores of three neighborhood social variables.

### 2.3 Analysis

After excluding participants with more than two missing items in CES-D and without available geocodes to merge the visibility factors (n=255), the analysis sample consisted of 1867 participants, in which 498 full twin pairs, 214 were monozygotic (MZ) and 284 dizygotic (DZ) (containing both same-sex and opposite-sex pairs), and 871 single twins (their cotwins were not included). Multi-PRS data was available in 702 individual twins. Zygosity was determined using DNA polymorphism analysis and/or a validated questionnaire. Missing values in the covariates were imputed using median values. Both self-reported CES-D score and visibility factors showed normal distributions by standardized normal probability plots.

#### 2.3.1 Environmental impact assessment

Linear multivariable regression models were first used to assess the relationship between visibility factors and self-reported depression, adjusting for all covariates. The clustering effect from twin pair sampling was accounted for by robust standard errors. To control for unobserved individual heterogeneity due to genetic and shared environmental factors, we conducted within-pair fixed-effects linear regression using the same covariates. This analysis was performed for all twin pairs and separately for MZ twins, where genetic influences are fully controlled.

To explore potential non-linear effects, generalized additive models (GAM) with penalized spline smoothing terms were used (Hastie & Tibshirani, 1986). Model tuning was performed through grid search, and the best-fitting models were identified based on the lowest Akaike Information Criterion (AIC; lower is better). Covariates were also included, and a random effect term was included to account for familial clustering. The linearity was tested by removing the smoothing term and comparing AIC values to determine whether the nonlinear model provided a better fit. The R package “mgcv” (version 1.9-1) was used (Wood, 2016).

The combined effect of visibility factors were assessed using quantile g-computation, which provided individual weights for both positive and negative associations (Keil et al., 2020). Covariates were included and the cluster effect was accounted for. The R package “qgcomp” (version 2.10.1) was used (Keil et al., 2020).

Because the analysis of variance showed significant sex-difference in the self-report CES-D score, sex-stratification was performed across all abovementioned models.

#### 2.3.2 Gene-environment correlation (rGE) analysis

Linear regression models between the multi-PRS score of depression and visibility factors were conducted to capture rGE related to depression. We did not adjust for covariates but applied robust standard error. Sex-stratification was performed.

Next, nonspecific rGE was explored using a twin design based on full twin pairs. We started with specification of a saturated twin model which examined the assumptions of equal means and variances by twin order and zygosity and tested sex-difference. Then, intrapair correlations (ρ) for visibility factors were calculated for MZ and DZ twins (all available individual twins) to decide to choose the full univariate ACE (including A, C, and E) or ADE (including A, D, and E) models, selecting the ADE model if the ρMZ was over twice the ρDZ (D is only explored when there is no evidence for C). Nested models (AE, E) were then fitted by sequentially dropping each variance component from the full model. The likelihood test between full and nested models and AIC were used to determine the best-fitting model (Røysamb & Tambs, 2016). Figure 1A presents the diagram of the example univariate AE model. Age was included as a covariate. The R package “OpenMx” (version 2.21.13) was used (Neale et al., 2016).

**Figure 1:**
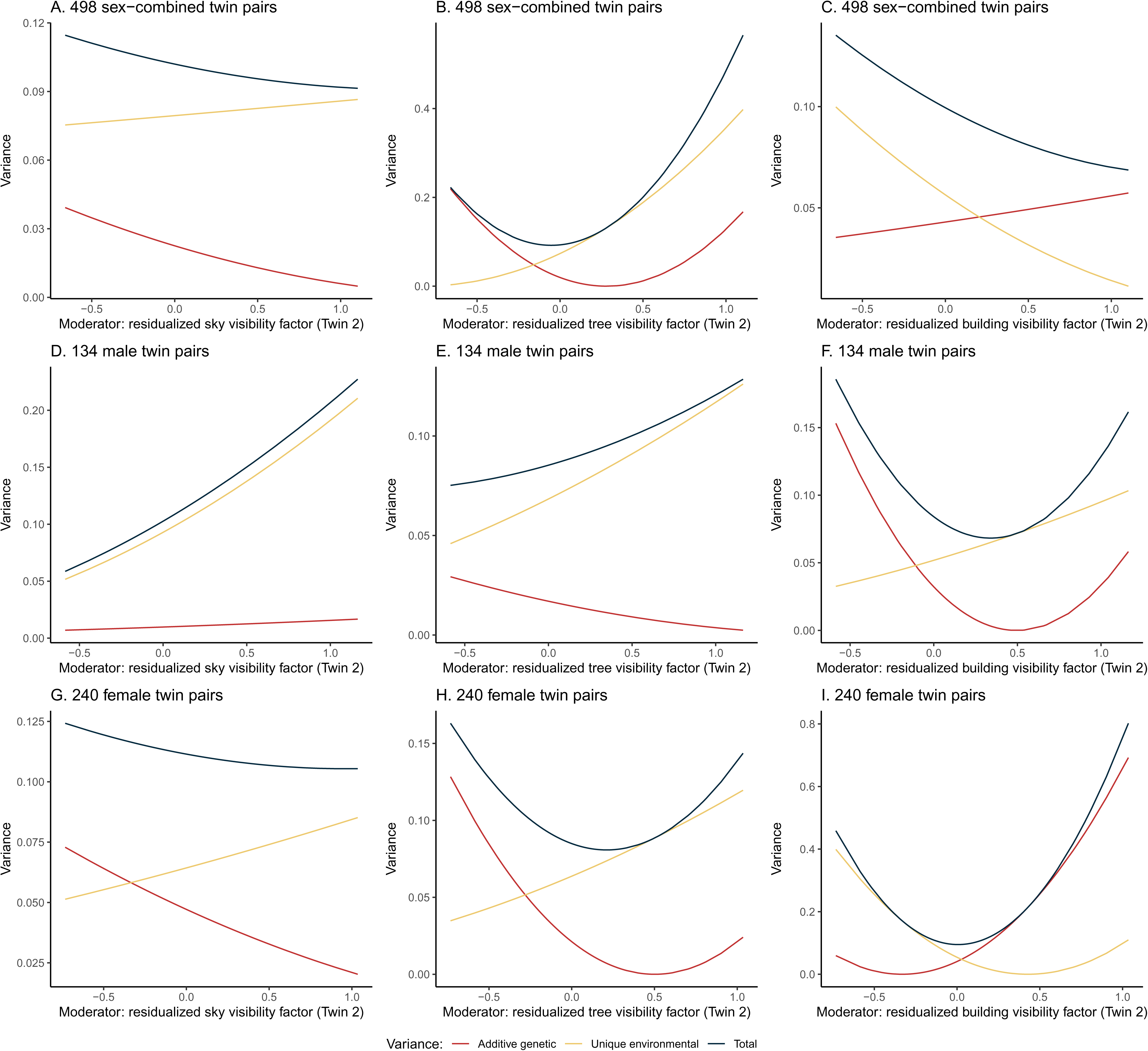
Diagrams of univariate (A) and bivariate moderation (B) AE twin model.

#### 2.3.3 Gene-environment interaction (G×E) analysis

First, we added the interaction term between visibility factors and the multi-PRS score of depression in the linear regression with the outcome of the self-report CES-D score. Sex-stratification was performed, covariates were adjusted for, and the robust standard error was applied.

Second, using the twin design, we applied the same procedure for the self-reported CES-D score to check assumptions, determine whether sex-stratification was necessary, and identify variance components to be included. Next, given our hypothesis that genetic factors contribute to the moderators (visibility factors), we employed a bivariate Cholesky moderation model to assess the moderation on the A, C/D, and E unique to the self-reported depression and those shared with the moderator (van der Sluis et al., 2012). In addition to standard paths for A, C/D, and E components, we introduced beta (β) terms to quantify the extent of moderation on both shared and unique paths of the self-report CES-D score (Purcell, 2002). These paths were further used to construct variance estimation for each component. Figure 1B presents an example AE moderation model. The self-report CES-D score and visibility factors were residualized by age as covariate adjustment before twin modeling. The R package “umx” (version 4.21.0) was used (Bates et al., 2019).

#### 2.3.4 Sensitivity analysis

First, linear regression models were fitted to assess the relationship between two comparison variables and self-reported depression. The same covariates were adjusted for, and robust standard errors were applied.

Second, to detect potential heterogenous effects by urbanicity, we obtained city center and shopping area boundary data from the Finnish Environment Institute (www.syke.fi) in 2019 to determine whether participants’ geocodes (used for generating visibility factors) were within urban areas. We conducted stratified linear regression analyses by urban residency, adjusting for covariates and applying robust standard errors.

## 3 Results

### 3.1 Characteristics of included participants

Table 1 presents the participants’ characteristics before imputation and their distributions between sexes. Among the 1867 participants (mean age: 37.2, standard deviation (SD): 1.5) included in the study, almost 60% were female. Most were employed (86%) and had received at least a bachelor-level education (65%). In their early midlife, 44% reported living with a spouse/partner and child(ren), 47% reported consuming alcohol monthly or less, 52% were never smokers, and 62% had never used illicit substances, such as marijuana. Regarding leisure-time physical activity, 32% of participants exercised at least 4–5 times per week, 35% were active 2–3 times per week, and 32% engaged in once a week or less. Univarite linear or ordinal logistic regression with robust standard error showed that most of these characteristics had a significant sex-difference except age, living status, and the deprivation score. The distributions of these characteristics between included participants and those available in the database were similar (Supplemental Table 2). The mean sky, tree, and building visibility factors were 0.6 (SD: 0.1), 0.3 (SD: 0.1), and 0.1 (SD: 0.1), respectively. The mean self-reported CES-D score was 1.9 (SD: 0.3) in sex-combined twins, 1.8 (SD: 0.3) in males, and 1.9 (SD: 0.3) in females. Supplemental Figure 1 presents the relationship between three visibility factors and the self-reported CES-D scores, while no obvious gradient or cluster was observed.

**Table 1:** Characteristic of included participants before imputation (individual twin n=1867)

| Characteristics | Missing | Sex-combined<br>(individual<br>twin n=1867) | Male<br>(individual<br>twin n=768) | Female<br>(individual<br>twin n=1099) | P-value for<br>sex-difference <sup>a</sup> |
| --- | --- | --- | --- | --- | --- |
| *Mean (standard deviation)* |  |  |  |  |  |
| **CES-D score** | 0 | 1.9 (0.3) | 1.8 (0.3) | 1.9 (0.3) | <0.01 |
| **Age (years)** | 0 | 37.2 (1.5) | 37.2 (1.4) | 37.2 (1.5) | 0.62 |
| **Deprivation score** | 0 | -0.0 (0.8) | 0.2 (0.8) | -0.01 (0.8) | 0.42 |
|  |  | *N. (%)* |  |  |  |
| **Work** | 2 |  |  |  | <0.01 |
| Not working or other situation |  | 262 (14.1) | 66 (8.6) | 196 (17.9) |  |
| Currently working |  | 1603 (86.0) | 702 (91.4) | 901 (82.1) |  |
| **Education** | 3 |  |  |  | <0.01 |
| Post-secondary or lower |  | 655 (35.1) | 333 (43.4) | 322 (29.4) |  |
| Bachelor/equivalent or above |  | 1209 (64.9) | 435 (56.6) | 774 (70.6) |  |
| **Living status** | 2 |  |  |  | 0.14 |
| A spouse/partner |  | 510 (27.4) | 225 (29.3) | 285 (26.0) |  |
| A spouse/partner and child(ren) |  | 827 (44.3) | 324 (42.2) | 503 (45.9) |  |
| Alone |  | 378 (20.3) | 185 (24.1) | 193 (17.6) |  |
| Other |  | 150 (8.0) | 34 (4.4) | 116 (10.6) |  |
| **Illicit substance use** | 2 |  |  |  | <0.01 |
| Never |  | 1164 (62.4) | 417 (54.4) | 747 (68.0) |  |
| A least once |  | 701 (37.6) | 350 (45.6) | 351 (32.0) |  |
| **Ever smoker (smoked over 100 cigarettes lifetime)** | 3 |  |  |  | <0.01 |
| Never |  | 974 (52.3) | 365 (47.7) | 609 (55.5) |  |
| Ever smoker |  | 890 (47.8) | 401 (52.4) | 489 (44.5) |  |
| **Alcohol** | 52 |  |  |  | <0.01 |
| At most monthly |  | 853 (47.0) | 258 (34.7) | 595 (55.6) |  |
| 2-4 times a month |  | 681 (37.5) | 305 (41.0) | 376 (35.1) |  |
| 2-3 times a week or more |  | 281 (15.5) | 181 (24.3) | 100 (9.3) |  |
| **Physical activity** | 1 |  |  |  | 0.02 |
| Once a week or less |  | 607 (32.5) | 263 (34.3) | 344 (31.3) |  |
| 2-3 times a week |  | 659 (35.3) | 284 (37) | 375 (34.1) |  |
| 4-5 times a week or more |  | 600 (32.2) | 220 (28.7) | 380 (34.6) |  |

<sup>a</sup> P-values were generated by linear or ordinal logistic regression with robust standard error (adjusted for the twin cohort's design effect) for sex-difference.

### 3.2 Association between visibility factors and self-reported depression

After adjusting for covariates, a higher level of building visibility factor was significantly associated with a higher self-reported CES-D score (coefficient: 0.20, 95% confidence interval (CI): 0.03, 0.36) in males (Table 2). The power for this multiple linear regression was 1. No significant associations appeared in models among females or when the sexes were combined. In within-pair linear regression models (all individual twins or MZ twins only), associations between any visibility factor and self-reported CES-D score were null, regardless of sex (Table 2).

**Table 2:** Relationship between visibility factors and self-reported depression using linear regression or within-pair fixed-effect regression.

| Visibility factor | Adjusted Coefficient (95% confidence interval) <sup>a</sup> |  |  |
| --- | --- | --- | --- |
|  | Linear regression for all twins | Within-pair linear regression for all twins | Within-pair linear regression for MZ twins only <sup>b</sup> |
| *Sex-combined (individual twin n=1867)* |  |  |  |
| Sky | 0.01 (-0.11, 0.14) | 0.03 (-0.25, 0.31) | 0.38 (-0.04, 0.81) |
| Tree | -0.07 (-0.19, 0.05) | 0.09 (-0.14, 0.31) | -0.07 (-0.44, 0.3) |
| Building | 0.07 (-0.06, 0.19) | -0.15 (-0.43, 0.13) | -0.27 (-0.71, 0.16) |
| *Male (individual twin n=768)* |  |  |  |
| Sky | -0.04 (-0.22, 0.13) | 0.33 (-0.24, 0.89) | 0.39 (-0.34, 1.12) |
| Tree | -0.15 (-0.31, 0.02) | -0.23 (-0.69, 0.24) | -0.00 (-0.67, 0.66) |
| Building | 0.20 (0.03, 0.36)* | -0.03 (-0.59, 0.52) | -0.41 (-1.21, 0.39) |
| *Female (individual twin n=1099)* |  |  |  |
| Sky | 0.05 (-0.12, 0.22) | 0.01 (-0.38, 0.40) | 0.39 (-0.13, 0.92) |
| Tree | -0.02 (-0.19, 0.15) | 0.16 (-0.15, 0.47) | -0.01 (-0.47, 0.45) |
| Building | -0.02 (-0.20, 0.15) | -0.24 (-0.64, 0.15) | -0.32 (-0.84, 0.21) |

\* P<0.05
<sup>a</sup> Adjusted for age, sex, education, work, living status, smoking, alcohol drinking, illicit substance use, physical activity, and neighborhood deprivation score. Adjusted for twin relatedness within pairs.
<sup>b</sup> There was a total of 669 MZ individual twins (263 were males and 406 were females).

No significant mixture effect of visibility factors on self-reported CES-D score was observed in sex-combined, male or female participants. No significant nonlinear relationship was detected, as well.

### 3.3 Results of rGE

No significant association was found between the multi-PRS score and visibility factors, regardless of sex (Supplemental Table 3).

There was a total of 498 full twin pairs. Assumptions of twin modeling were generally met. No significant sex-difference in visibility factors was detected (Supplemental Table 4). There was no following sex-stratification for univariate modeling of visibility factors. The ratios between ρMZ and ρDZ were below 2 for the sky and building visibility factors (the ACE model correspondingly), and the ratio was over 14 for the tree visibility factor (the ADE model correspondingly) (Supplemental Table 5). The AE models for both sky and building visibility factors had the lowest AIC and were identified as the most optimal (Table 3). Although the ADE model for the tree visibility factor had a lower AIC than the AE model, we still decided to identify the AE model as the most optimal, given a small difference between AICs of two models (<2) and negative estimation of A in the ADE model (Table 3). The additive genetic component (A) contributed to 46% (CI: 0.37, 0.55), 29% (CI: 0.17, 0.39), and 49% (CI: 0.40, 0.57) to the total variances of sky, tree, and building visibility factors, respectively (Table 3).

**Table 3:** Estimates of variance components in univariate twin modelling for visibility factors (twin pair n=498)

| Visibility factor | Model | Standardized variance (95% confidence interval) |  |  |  | AIC | P-value <sup>a</sup> |
| --- | --- | --- | --- | --- | --- | --- | --- |
|  |  | A (additive genetic) | C (common environmental) | D (dominant genetic) | E (unique environmental) |  |  |
| Sky | ACE | 0.26 (-0.03, 0.54) | 0.18 (-0.06, 0.40) | / | 0.56 (0.47, 0.67) | -1617.06 | Ref. |
|  | AE | 0.46 (0.37, 0.55) | / | / | 0.54 (0.45, 0.63) | -1616.88 | 0.14 |
|  | E | / | / | / | 1 | -1547.22 | <0.01 |
| Tree | ADE | -0.10 (-0.58, 0.36) | / | 0.44 (-0.07, 0.96) | 0.67 (0.56, 0.79) | -1438.55 | Ref. |
|  | AE | 0.29 (0.17, 0.39) | / | / | 0.71 (0.61, 0.83) | -1437.74 | 0.09 |
|  | E | / | / | / | 1 | -1416.69 | <0.01 |
| Building | ACE | 0.26 (-0.02, 0.54) | 0.20 (-0.03, 0.41) | / | 0.54 (0.45, 0.65) | -1577.65 | Ref. |
|  | AE | 0.49 (0.40, 0.57) | / | / | 0.51 (0.43, 0.60) | -1576.76 | 0.09 |
|  | E | / | / | / | 1 | -1500.38 | <0.01 |

<sup>a</sup> P-values were generated by the likelihood tests for model comparison.

### 3.4 Results of G×E

Based on linear regression, the effects of interaction terms between visibility factors and the multi-PRS score on the self-reported CES-D score were insignificant, regardless of sex (Supplemental Table 6).

The saturated model indicated a significant sex difference in the self-reported CES-D score (Supplemental Table 4), and ρMZ and ρDZ indicated to use the full ACE initially, regardless of sex (Supplemental Table 7). Univariate twin models identified AE as the most optimal for models among sex-combined (498 twin pairs), male (134 twin pairs), and female pairs (240 twin pairs) (Supplemental Table 8). The moderation model was conducted separately for male, female, and sex-combined twin pairs, excluding opposite-sex pairs. Results are presented in Figure 2 (unstandardized variance estimates), Supplemental Figure 2 (standardized variance estimates), and Supplemental Table 9 (path coefficients and β estimates). In most models, lines of A and E were non-parallel and often intersected, except for the model with the moderator of building visibility factor in male twin pairs (Figure 2D). In sex-combined twin pairs, lines intersected by tree and building visibility factors (Figures 2B and 2C). In male twin pairs, this intersection occurred by the building visibility factor (Figure 2F), while in female twin pairs, intersections were observed by all three visibility factors (Figures 2G, 2H, and 2I). Notably, with the moderator of the building visibility factor, variances contributed by A initially exceeded variances contributed by E in males when the building visibility factor was very low, whereas the opposite pattern was observed in females.

**Figure 2:**
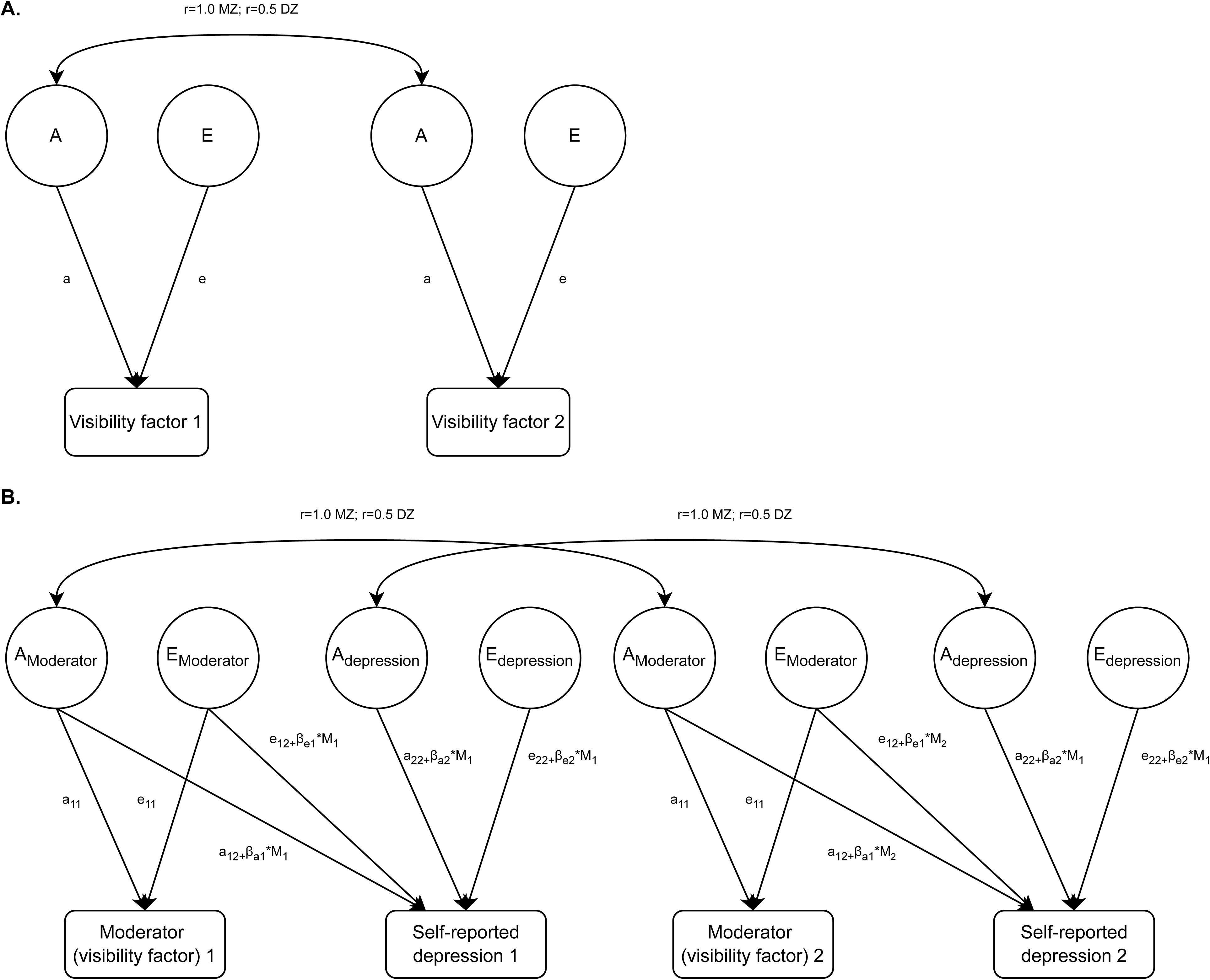
Unstandardized variance estimation of additive genetic and unique environmental components based on bivariate AE moderation twin models.

### 3.5 Sensitivity analysis

There was no significant association between the mean tree cover density and self-reported CES-D score (Supplemental Table 10). However, a higher percentage of impervious areas within a 100 m buffer was significantly associated with a higher self-reported CES-D score (coefficient: 0.11, 95% CI: 0.02, 0.21) in males. In the unadjusted linear regression among males, the variance of the self-reported CES-D score was explained 0.5% by the building visibility factors and 0.7% by the percentage of impervious areas within a 100 m buffer.

A quarter of participants (25.5%) lived in urban areas as defined above. Regardless of urban residency, there was no significant association between visibility factors and self-reported CES-D score among sex-combined, male, or female twins (Supplemental Table 11).

## 4 Discussion

Based on nearly two thousand participants in Finland, we investigated the environmental impact of the visibility of the environment (sky, tree, and building visibility factors) at the residence on self-reported symptoms of depression in early midlife and studied the gene-environment interplay through both molecular method and twin design. In males, a higher level of building visibility factor was associated with a higher depression score, while the association attenuated to null after controlling for individual unobserved heterogeneity (genetics, etc.). Univariate twin models indicated that additive genetic effect variance contributed substantially to the variance of all three visibility factors, with nearly 50% of the variance of tree and building visibility factors, as rGE. Bivariate twin moderation models showed a strong G×E between visibility factors and self-reported depression.

Our findings partially align with previous research that connect the visibility of building in the living environment with self-reported depression in males, which possibly due to the obstruction of an ideal view. A Norwegian longitudinal quasi-experiment involving nearly 300 coronary and pulmonary patients found that rooms with blocked views by city center or buildings in the rehabilitation center negatively impacted the change of mental health in males, whereas no such effect was observed in females (Raanaas et al., 2011). Yao et al. (2024a) examined all three visibility factors from windows among 160 participants and found that a higher ratio of building visibility factor aggravated the negative effects on the beta indicator of brain waves, reflecting stress and the inability to relax. They further summarized that the positive effect of building elements in window views on mental health is limited (Yao et al., 2024b). Sensitivity analysis using satellite-based measures of built-up areas yielded similar results. A longitudinal cohort study in Hong Kong, China found that increased building-block density was associated with a higher odd of depressive symptoms (Sarkar et al., 2021), while a Danish case-control study suggested the relationship may be nonlinear and influenced by socioeconomic status (Chen et al., 2025). We did not observe any salient effect from other visibility factors, despite prior research demonstrating the favorable effect from the visibility of sky and greenspaces (Hou et al., 2024b; Shu et al., 2022). One possible explanation is that our visibility factors may not accurately reflect the actual view from individual rooms in which the participants reside.

A notable genetic predisposition was observed in the visibility of the environment around residences during participants’ early midlife, suggesting that heritable traits of parents influence the living environments their offspring choose after leaving the parental home and, perhaps, several times of moving. For instance, migrant workers in China often settle in midsize or large cities near their hometowns for economic and career reasons (Jinqi et al., 2019) (larger cities usually have higher building density), while in Europe and South America, social status can influence how urban landscapes are perceived and used, potentially shaping individuals’ residential decisions (de la Barrera et al., 2016; Riechers et al., 2018). An Israeli study identified socioeconomic status as one of the primary determinants of residential location choice and culture-oriented lifestyles (such as attending cinemas, museums, and concerts) as one of the substantial secondary determinants, among knowledge workers (Frenkel et al., 2013). Abundant cultural facilities are usually found in cities. Numerous twin studies have demonstrated genetic influences on these factors (Faßbender et al., 2019; Polderman et al., 2015), supporting the idea of rGE in choosing the living environment.

To our knowledge, no previous study has examined the visibility of the environment at the residence and mental health under the G×E context, but some have used satellite-based exposure measures. In the UK Biobank, a higher PRS for depression was associated with living in areas with less greenspace, and Mendelian randomization strengthened the causal inference (Reed et al., 2022). Another UK Biobank study found that participants with higher CRHR1 genetic risk who lived in areas with greater urban environmental exposure (including greenspace proximity and land use density) had more severe affective symptoms, including depression (Xu et al., 2023). The G×E effect may be explained by biological mechanisms, such as inflammation, and, as abovementioned, stress or urban temperature induced or reflected by the visibility of the environment were known as risk factors for inflammation (Rohleder, 2019; Wang et al., 2020). Gal et al. (2024)identified 23 genes linked to stress-related depression that are involved in inflammatory processes. A review highlighted inconsistencies in the immune effects of genetic variants on depression, potentially due to environmental influences or G×E (Barnes et al., 2017). The relationship between the visibility of the environment at the residence and depression is likely governed by a complicated cascade of biological mechanisms.

There are some limitations in our study, which need to be addressed. First is the sample size in the twin modeling. Due to the loss of follow-up, fewer complete twin pairs were available, and sex-stratification further reduced the sample size in each model, affecting the power to estimate path and β coefficients. Second, due to sex stratification, the multiple testing issue arose which increases the risk of type-1 error. Third, the visibility of the environment at the residence is a multifaceted concept. Natural features, visual harmony, spatial proportions, identity, and visual disturbances were identified in urban views, may exert heterogeneous effects on depression, as well as mental health (Karimimoshaver et al., 2020). Fourth, one could consider the assessment of photographs of the exact window views that individuals have at their residences to obtain a more adequate representation of the likely indoor exposure to view factors. Fifth, Finland is known for prevalent green spaces, even in urban areas generally, which may mitigate any negative effect of poor visibility factors from residential locations. There was a generalizability issue, and replication in other countries is encouraged. Sixth, visibility factors were assessed only at residential locations, but adults spend a large proportion of their time at work. Expanding exposure assessments to workplace could provide a more comprehensive understanding of environmental influences on mental health.

## 5. Conclusion

The findings of this study have implications for health-related urban design, suggesting that placing tower blocks farther apart, skyward dis-densification, and reducing neighborhood scale may help mitigate environmental risk for depression. Additionally, the evidence of gene-environment interplay informs public health strategies in reducing exposure for vulnerable populations, including those with genetic susceptibilities, and enhancing personalized interventions.

## Author statement

Z.W., M.P., S.K., and J.K. contributed to the study conception and design. M.P., S.A., J.S., D.K., S.K., and J.K. contributed to the data collection. Z.W. and S.Z. performed the data analysis, and Z.W. drafted the manuscript. All authors reviewed and approved the manuscript. J.K. provided critical supervision and guidance.

## Acknowledgement

Data collection in FinnTwin12 has been supported by the National Institute on Alcohol Abuse and Alcoholism (grants AA-12502, AA-00145, and AA-09203 to Richard J. Rose, and AA015416 to Danielle Dick and Jessica Salvatore) and the Academy of Finland (grants 100499, 205585, 118555, 141054, 264146, 308248, 312073, 336823, and 352792 to Jaakko Kaprio). Jaakko Kaprio acknowledges support by the Academy of Finland (grants 265240, 263278). S.K. was in part funded by the Funded by the European Union (ERC-2022-CoG-BrainScape-101086188). Views and opinions expressed are however those of the authors only and do not necessarily reflect those of the European Union or the European Research Council Executive Agency (ERCEA). Neither the European Union nor the granting authority can be held responsible for them.

We acknowledge the effort from the Equal-life scientific team for the exposure enrichment in the sensitivity analysis.

## Ethics statements

The ethics committee of the Department of Public Health of the University of Helsinki (Helsinki, Finland) and the Institutional Review Board of Indiana University (Bloomington, Indiana, USA) approved the FinnTwin12 study protocol from the start of the cohort. The ethical approval of the ethics committee of the Helsinki University Central Hospital District (HUS) is the most recent and covers the most recent data collection (early midlife) (HUS/2226/2021, dated September 22, 2021). All participants and their parents/legal guardians gave informed written consent to participate in the study. The authors assert that all procedures contributing to this work comply with the ethical standards of the relevant national and institutional committees on human experimentation and with the Helsinki Declaration of 1975, as revised in 2008.

## Data availability statement

The FinnTwin12 data are not publicly available due to the restrictions of informed consent. However, the FinnTwin12 data are available through the Institute for Molecular Medicine Finland (FIMM) Data Access Committee (DAC) for authorized researchers who have IRB/ethics approval and an institutionally approved study plan. To ensure the protection of privacy and compliance with national data protection legislation, a data use/transfer agreement is needed, the content and specific clauses of which will depend on the nature of the requested data. Requests will be addressed in a reasonable time frame (generally two to three weeks), and the primary mode of data access is by either personal visit or remote access to a secure server. Codes for major analyses are available at https://github.com/doge73/viewfactor_dep).

