## Supplemental Figure 1-3 and Table 1-11 for "Gene-environment interplay in the relationship between the visibility of the environment and self-reported depression in early midlife: a Finnish twin cohort study"

Zhyang Wang et al. Online supplemental material

Table of content

Supplemental Figure 1: Ternary plots between visibility factors and CES-D scores among sex-combined (A), male (B), and female (C) individual twins

Supplemental Figure 2: Standardized variance estimation of additive genetic and unique environmental components based on bivariate moderation models

Supplemental Table 1: The description of polygenic risk scores (PRSes)

Supplemental Table 2: Characteristics of included participants and those available in the database

Supplemental Table 3: The relationship between multi-PRS score and visibility factors using linear regression

Supplemental Table 4: Saturated models for assumption testing of univariate twin modeling and detecting sex-difference

Supplemental Table 5: Intrapair correlation for visibility factors (individual twin n=1867)

Supplemental Table 6: The effect of interaction between the multi-PRS score and visibility factors on self-reported CES-D score using linear regression

Supplemental Table 7: Intrapair correlation for the self-reported CES-D score

Supplemental Table 8: Estimates of variance components in univariate twin modelling for the self-reported CES-D score

Supplemental Table 9: Path coefficient and β estimation based on bivariate moderation models

Supplemental Table 10: The relationship between comparison factors and the self-reported CES-D score using linear regression

Supplemental Table 11: The relationship between visibility factors and the self-reported CES-D score stratified by living in urban areas or not using linear regression

Supplemental Figure 1: Ternary plots between visibility factors and CES-D scores among sex-combined (A), male (B), and female (C) individual twins

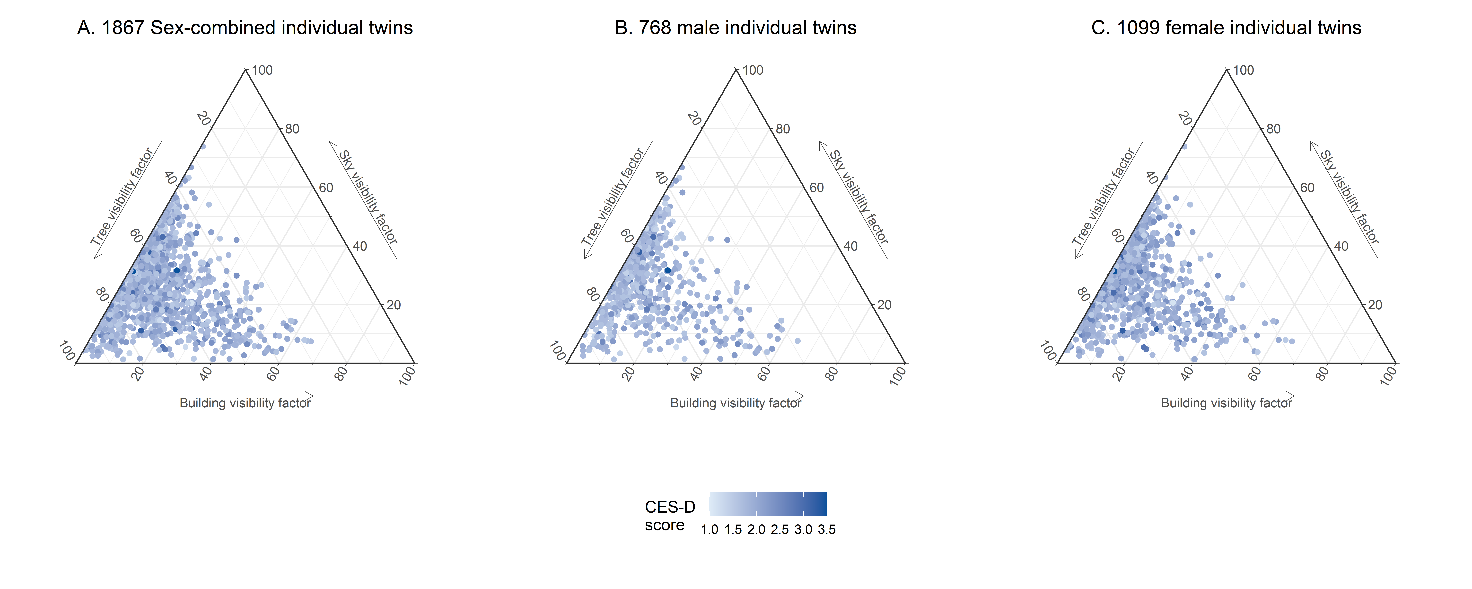

Supplemental Figure 2: Standardized variance estimation of additive genetic and unique environmental components based on bivariate moderation models

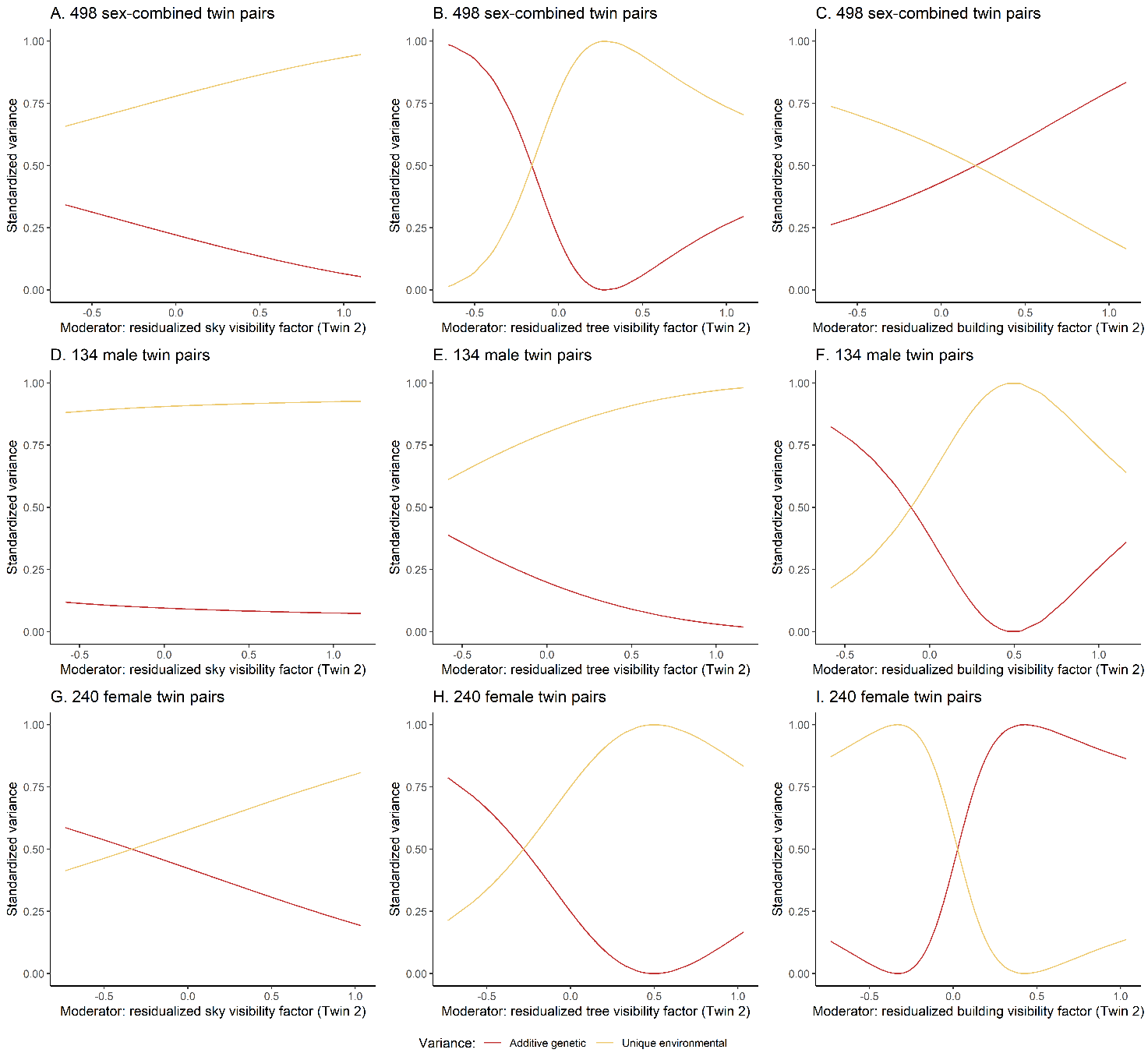

Supplemental Table 1: The description of polygenic risk scores (PRSes)

| PRS for phenotype | Heritability | Used tool | Source study (PMID) | Source sample size | Source phenotype ascertainment |
| --- | --- | --- | --- | --- | --- |
| Schizophrenia | 0.24 | LDpred+PLINK | 25056061 | 36,989 cases and 113,075 controls | Individuals with schizophrenia or schizoaffective disorder |
| Body mass index | 0.17 | LDpred+PLINK | 30124842 | 681,275 participants | / |
| Alzheimer’s disease | 0.10 | LDpred+PLINK | 24162737 | 17,008 cases and 37,154 controls | Varied by cohorts (autopsy based on DSM-IV, NINCDS/ADRDA criteria, etc. ) |
| Neuroticism (raw score) | 0.09 | LDpred+PLINK | http://www.nealelab.is/uk-biobank/ | / | / |
| Broad depression (defined as having seen a doctor for nerves, anxiety, tension, or depression) | 0.06 | LDpred+PLINK | <http://www.nealelab.is/uk-biobank/> | / | / |
| Subjective well-being | 0.03 | LDpred+PLINK | 27089181 | 298420 participants | Questions varied by cohorts, usually the sum of positive affect and life satisfaction scores |
| Major depressive disorder | 0.07 | LDpred+PLINK | 29700475 | 135,458 cases and 344,901 controls | Structured diagnostic interviews, National inpatient electronic records, Structured diagnostic interview, Kaiser Permanente Northern California Healthcare electronic medical records (1995–2013), National inpatient electronic records, self-reported MDD symptoms or treatment or electronic records, or treatment for clinical depression by a medical professional |
| Insomnia | 0.04 | LDpred+PLINK | 30804565 | 1,331,010 participants | In UK Biobank, insomnia cases were defined as participants who answered this question with “usually”, while participants answering “never/rarely” or “sometimes” were defined as controls in question "“Do you have trouble falling asleep at night or do you wake up in the middle of the night?” A structured flow with several questions in 23andMe. |
| Alcohol dependence | 0.08 | LDpred+PLINK | 30482948 | 4,904 cases and 37,944 controls | Questionnaire varied by cohorts (Semi-Structured Assessment for the Genetics of Alcoholism, etc.) following DSM criteria |
| Externalizing problems | 0.06 | LDpred+PLINK | 34446935 | up to 1,251,809 participants | Consist of attention-deficit/hyperactivity disorder, problematic alcohol use, lifetime cannabis use, reverse-coded age at first sexual intercourse, number of sexual partners, general risk tolerance, and lifetime smoking initiation, by varied questionnaire. |
| Smoking initiation | 0.09 | LDpred+PLINK | 36477530 | 3,383,199 participants | Binary phenotype with any participant reporting ever being a regular smoker in their life (current or former) coded “2”, while any participant who reported never being a regular smoker in their life coded “1”. Does not include information about pipes/cigar/chew, or other non-cigarette forms of tobacco use. This phenotype was measured in a variety of ways. a. Have you smoked over 100 cigarettes over the course of your life? b. Have you ever smoked every day for at least a month? c. Have you ever smoked regularly? |
| Educational attainment | 0.10 | LDpred+PLINK | 30038396 | 1,131,881 participants | ISCED classification |

Supplemental Table 2: Characteristics of included participants and those available in the database

| **Characteristics** | **Included in study** | | **Available in the database** | |
| --- | --- | --- | --- | --- |
|  | Missing | N. (%) / Mean (SD) | Missing | N. (%) / Mean (SD) |
| **Age (years)** | 0 | 37.2 (1.5) | 0 | 37.2 (1.5) |
| **Deprivation score** | 0 | -0.0 (0.8) | 196 | 0.00 (0.8) |
| **Sex** | 0 |  | 0 |  |
| Male |  | 768 (41.1) |  | 876 (41.3) |
| Female |  | 1099 (58.9) |  | 1246 (58.7) |
| **Work** | 2 |  | 18 |  |
| Not working or other situation |  | 262 (14.1) |  | 297 (14.1) |
| Currently work |  | 1603 (86.0) |  | 1807 (85.9) |
| **Education** | 3 |  | 18 |  |
| Post-secondary or lower |  | 655 (35.1) |  | 760 (36.1) |
| Bachelor/equivalent or above |  | 1209 (64.9) |  | 1344 (63.9) |
| **Living status** | 2 |  | 15 |  |
| A spouse/partner |  | 510 (27.4) |  | 583 (27.7) |
| A spouse/partner and child(ren) |  | 827 (44.3) |  | 958 (45.5) |
| Alone |  | 378 (20.3) |  | 397 (18.8) |
| Other |  | 150 (8.0) |  | 169 (8.0) |
| **Illicit substance use** | 2 |  | 55 |  |
| Never |  | 1164 (62.4) |  | 1318 (63.8) |
| A least once |  | 701 (37.6) |  | 749 (36.2) |
| **Ever smoker (smoked over 100 cigarettes lifetime)** | 3 |  | 57 |  |
| Never |  | 974 (52.3) |  | 1082 (52.4) |
| Ever smoker |  | 890 (47.8) |  | 983 (47.6) |
| **Alcohol** | 52 |  | 112 |  |
| At most monthly |  | 853 (47.0) |  | 927 (46.1) |
| 2-4 times a month |  | 681 (37.5) |  | 760 (37.8) |
| 2-3 times a week or more |  | 281 (15.5) |  | 323 (16.1) |
| **Physical activity** | 1 |  | 33 |  |
| Once a week or less |  | 607 (32.5) |  | 700 (33.5) |
| 2–3 times a week |  | 659 (35.3) |  | 730 (34.9) |
| 4–5 times a week or more |  | 600 (32.2) |  | 659 (31.6) |

N: number of participants; SD: standard deviation

Supplemental Table 3: The relationship between multi-PRS score and visibility factors using linear regression

| **Outcome: visibility factor** | **Coefficient (95% CI)** | | |
| --- | --- | --- | --- |
|  | Sex-combined (individual twin n=702) | Male (individual twin n=291) | Female (individual twin n=411) |
| Sky | -0.01 (-0.1, 0.08) | -0.11 (-0.31, 0.09) | 0.06 (-0.11, 0.23) |
| Tree | 0.06 (-0.04, 0.15) | 0.08 (-0.13, 0.29) | -0.03 (-0.20, 0.15) |
| Building | -0.04 (-0.13, 0.06) | 0.04 (-0.16, 0.25) | -0.03 (-0.19, 0.14) |

CI: confidence interval

Supplemental Table 4: Saturated models for assumption testing of univariate twin modeling and detecting sex-difference

| **Variable** | **Model** | **Estimated parameters** | **-2 loglikelihood  (-2ll)** | **Degree of freedom (df)** | **AIC** | **Difference of -2ll** | **Difference of df** | **Two-sided P value ^a^** |
| --- | --- | --- | --- | --- | --- | --- | --- | --- |
| Sky | Saturated | 25 | 2784.27 | 971 | 2834.27 | Ref. | Ref. | Ref. |
|  | Constrain expected Means to be equal across twin order | 21 | 5964.96 | 975 | 6006.96 | 3180.69 | 4 | <0.01 |
|  | Constrain expected Means and Variances to be equal across twin order | 17 | -1637.54 | 979 | -1603.54 | -4421.81 | 8 | 1 |
|  | Constrain expected Means and Variances to be equal across twin order and zygosity | 13 | -1635.38 | 983 | -1609.38 | -4419.65 | 12 | 1 |
|  | Constrain expected Means and Variances to be equal across twin order and zygosity and pairs' same/opposite sex | 9 | 1683.76 | 987 | 1701.76 | -1100.51 | 16 | 1 |
|  | Constrain expected Means and Variances to be equal across twin order and zygosity and pairs' same/opposite sex and sex | 7 | -1631.35 | 989 | -1617.35 | -4415.62 | 18 | 1 |
| Tree | Saturated | 25 | -1462.60 | 971 | -1412.60 | Ref. | Ref. | Ref. |
|  | Constrain expected Means to be equal across twin order | 21 | -1453.51 | 975 | -1411.51 | 9.09 | 4 | 0.06 |
|  | Constrain expected Means and Variances to be equal across twin order | 17 | -1451.84 | 979 | -1417.84 | 10.76 | 8 | 0.22 |
|  | Constrain expected Means and Variances to be equal across twin order and zygosity | 13 | -1451.50 | 983 | -1425.50 | 11.10 | 12 | 0.52 |
|  | Constrain expected Means and Variances to be equal across twin order and zygosity and pairs' same/opposite sex | 9 | -1450.46 | 987 | -1432.46 | 12.14 | 16 | 0.73 |
|  | Constrain expected Means and Variances to be equal across twin order and zygosity and pairs' same/opposite sex and sex | 7 | -1449.81 | 989 | -1435.81 | 12.79 | 18 | 0.80 |
| Building | Saturated | 25 | 1831.63 | 971 | 1881.63 | Ref. | Ref. | Ref. |
|  | Constrain expected Means to be equal across twin order | 21 | 7771.54 | 975 | 7813.54 | 5939.92 | 4 | <0.01 |
|  | Constrain expected Means and Variances to be equal across twin order | 17 | -1602.18 | 979 | -1568.18 | -3433.81 | 8 | 1.00 |
|  | Constrain expected Means and Variances to be equal across twin order and zygosity | 13 | -1601.66 | 983 | -1575.66 | -3433.29 | 12 | 1.00 |
|  | Constrain expected Means and Variances to be equal across twin order and zygosity and pairs' same/opposite sex | 9 | 1749.55 | 987 | 1767.55 | -82.07 | 16 | 1.00 |
|  | Constrain expected Means and Variances to be equal across twin order and zygosity and pairs' same/opposite sex and sex | 7.00 | -1597.62 | 989.00 | -1583.62 | -3429.25 | 18.00 | 1.00 |
| Self-reported CES-D score | Saturated | 25.00 | 441.40 | 971.00 | 491.40 | Ref. | Ref. | Ref. |
|  | Constrain expected Means to be equal across twin order | 21.00 | 443.44 | 975.00 | 485.44 | 2.04 | 4.00 | 0.73 |
|  | Constrain expected Means and Variances to be equal across twin order | 17.00 | 448.39 | 979.00 | 482.39 | 6.99 | 8.00 | 0.54 |
|  | Constrain expected Means and Variances to be equal across twin order and zygosity | 13.00 | 463.19 | 983.00 | 489.19 | 21.79 | 12.00 | 0.04 |
|  | Constrain expected Means and Variances to be equal across twin order and zygosity and pairs' same/opposite sex | 9.00 | 475.70 | 987.00 | 493.70 | 34.30 | 16.00 | <0.01 |
|  | Constrain expected Means and Variances to be equal across twin order and zygosity and pairs' same/opposite sex and sex | 7.00 | 482.76 | 989.00 | 496.76 | 41.36 | 18.00 | <0.01 |

^a^ P-value was from likelihood tests between different models.

Supplemental Table 5: Intrapair correlation for visibility factors (individual twin n=1867)

| **Visibility factor** | **Intrapair correlation (ρ)** | | **ρMZ/ρDZ ratio** |
| --- | --- | --- | --- |
|  | Monozygotic (MZ) | Dizygotic (DZ) |  |
| Sky | 0.44 (0.33, 0.54) | 0.31 (0.21, 0.42) | 1.39 |
| Tree | 0.32 (0.20, 0.45) | 0.02 (0.00, 0.16) | 14.46 |
| Building | 0.45 (0.35, 0.56) | 0.30 (0.19, 0.41) | 1.51 |

Supplemental Table 6: The effect of interaction between the multi-PRS score and visibility factors on self-reported CES-D score using linear regression

| **Interaction term between the multi-PRS score and visibility factors:** | **Coefficient (95% CI) ^a^** | | |
| --- | --- | --- | --- |
|  | Sex-combined (individual twin n=702) | Male (individual twin n=291) | Female (individual twin n=411) |
| Sky | 1.07 (-1.27, 3.41) | -0.49 (-5.77, 4.79) | 1.95 (-2.85, 6.74) |
| Tree | -0.47 (-2.74, 1.81) | 1.28 (-3.49, 6.05) | -1.29 (-6.24, 3.65) |
| Building | -0.38 (-2.93, 2.17) | -1.09 (-6.45, 4.27) | -0.58 (-5.43, 4.28) |

^a^ Adjusted for age, sex, education, work, living status, smoking, alcohol drinking, illicit substance use, physical activity, and neighborhood deprivation score

CI: confidence interval

Supplemental Table 7: Intrapair correlation for the self-reported CES-D score

| **Self-reported CES-D score** | **Intrapair correlation (ρ)** | | **ρMZ/ρDZ ratio** |
| --- | --- | --- | --- |
|  | Monozygotic (MZ) | Dizygotic (DZ) |  |
| Sex-combined  (individual twin n=702) | 0.28 (0.15, 0.41) | 0.16 (0.03, 0.30) | 1.74 |
| Male  (individual twin n=291) | 0.06 (0.00, 0.32) | 0.08 (0.00, 0.41) | 0.80 |
| Female  (individual twin n=411) | 0.32 (0.17, 0.47) | 0.44 (0.27, 0.61) | 0.72 |

Supplemental Table 8: Estimates of variance components in univariate twin modelling for the self-reported CES-D score

| **Self-reported CES-D score** | **Model** | **Standardized variance (95% CI)** | | | **AIC** | **P-value ^a^** |
| --- | --- | --- | --- | --- | --- | --- |
|  |  | A (additive  genetic) | C (common environmental) | E (unique environmental) |  |  |
| Sex-combined (twin pair n=498) | ACE | 0.25 (-0.09, 0.58) | 0.02 (-0.23, 0.27) | 0.72 (0.60, 0.86) | 499.33 | Ref. |
|  | AE | 0.28 (0.17, 0.39) | / | 0.72 (0.61, 0.83) | 497.36 | 0.85 |
|  | E | / | / | 1 | 517.57 | <0.01 |
| Male (twin pair n=134) | ACE | -0.19 (-0.93, 0.52) | 0.28 (-0.22, 0.78) | 0.91 (0.63, 1.22) | 131.07 | Ref. |
|  | AE | 0.20 (-0.05, 0.41) | / | 0.80 (0.59, 1.05) | 130.28 | 0.27 |
|  | E | / | / | 1 | 130.80 | 0.15 |
| Female (twin pair n=240) | ACE | -0.07 (-0.52, 0.42) | 0.35 (-0.08, 0.71) | 0.72 (0.59, 0.87) | 196.06 | Ref. |
|  | AE | 0.31 (0.17, 0.43) | / | 0.69 (0.57, 0.83) | 196.64 | 0.11 |
|  | E | / | / | 1 | 213.29 | <0.01 |

^a^ P-values were generated by the likelihood tests for model comparison.

AIC: Akaike Information Criterion; CI: confidence interval

Supplemental Table 9: Path coefficient and β estimation based on bivariate moderation models

| **Visibility factor** | **Path or β** | **Coefficient (95% confidence interval)** | | |
| --- | --- | --- | --- | --- |
|  |  | Sex-combined (twin pair n=498) | Male (twin pair n=134) | Female (twin pair n=240) |
| Sky | a_11_ | 0.08 (0.07, 0.08) | 0.08 (0.07, 0.10) | 0.00 (0.00, 0.01) |
|  | e_11_ | 0.08 (0.07, 0.09) | 0.08 (0.07, 0.09) | 0.11 |
|  | a_21_ | -0.02 (-0.06, 0.02) | -0.03 (-0.10, 0.03) | 0.16 |
|  | e_21_ | 0.02 (-0.01, 0.05) | 0.03 (-0.03, 0.09) | 0.01 |
|  | a_22_ | 0.17 (0.13, 0.20) | 0.13 (0.07, 0.20) | 0.06 |
|  | e_22_ | 0.26 (0.24, 0.29) | 0.27 (0.23, 0.32) | 0.25 |
|  | β_a1_ | 0.00 (-0.26, 0.26) | -0.06 (-0.46, 0.36) | -0.05 (-1.12, 1.32) |
|  | β_e1_ | -0.04 (-0.27, 0.20) | 0.01 (-0.40, 0.41) | -0.03 (-1.02, 0.87) |
|  | β_a2_ | -0.07 (-0.40, 0.26) | 0.09 (-0.58, 0.63) | -0.02 |
|  | β_e2_ | 0.05 (-0.16, 0.25) | 0.13 (-0.18, 0.41) | 0.07 (-0.76, 0.82) |
| Tree | a_11_ | 0.06 (0.05, 0.08) | 0.08 (0.05, 0.10) | 0.06 (0.05, 0.07) |
|  | e_11_ | 0.10 (0.09, 0.11) | 0.10 (0.09, 0.11) | 0.10 (0.09, 0.11) |
|  | a_21_ | -0.02 (-0.07, 0.03) | -0.01 (-0.09, 0.07) | -0.02 (-0.08, 0.05) |
|  | e_21_ | 0.01 (-0.02, 0.04) | -0.01 (-0.07, 0.05) | 0.00 (-0.03, 0.04) |
|  | a_22_ | 0.16 (0.12, 0.19) | 0.14 (0.08, 0.21) | 0.16 (0.12, 0.20) |
|  | e_22_ | 0.26 (0.24, 0.28) | 0.27 | 0.25 (0.22, 0.27) |
|  | β_a1_ | -0.41 (-0.75, -0.02) | -0.3 (-0.83, 0.19) | 0.00 (-0.43, 0.46) |
|  | β_e1_ | 0.21 (-0.07, 0.45) | 0.14 (-0.31, 0.58) | -0.01 (-0.32, 0.30) |
|  | β_a2_ | -0.09 (-0.47, 0.31) | 0.23 (-0.50, 0.86) | -0.29 (-0.64, 0.05) |
|  | β_e2_ | 0.12 (-0.11, 0.34) | -0.06 (-0.34, 0.29) | 0.10 (-0.14, 0.33) |
| Building | a_11_ | 0.08 (0.07, 0.09) | 0.09 (0.07, 0.10) | 0.07 (0.06, 0.09) |
|  | e_11_ | 0.08 (0.07, 0.09) | 0.08 (0.07, 0.09) | 0.08 (0.07, 0.09) |
|  | a_21_ | 0.04 (0.00, 0.09) | 0.06 (-0.03, 0.15) | 0.03 (-0.02, 0.09) |
|  | e_21_ | -0.02 (-0.06, 0.01) | -0.04 (-0.12, 0.04) | -0.00 (-0.05, 0.04) |
|  | a_22_ | 0.16 (0.12, 0.20) | 0.12 (0.05, 0.19) | 0.17 (0.12, 0.20) |
|  | e_22_ | 0.26 (0.24, 0.28) | 0.27 (0.23, 0.32) | 0.24 (0.21, 0.26) |
|  | β_a1_ | -0.15 (-0.39, 0.10) | -0.20 (-0.53, 0.13) | 0.02 (-0.39, 0.45) |
|  | β_e1_ | 0.07 (-0.16, 0.31) | 0.19 (-0.13, 0.51) | -0.15 (-0.43, 0.16) |
|  | β_a2_ | 0.18 (-0.21, 0.50) | -0.16 | 0.58 (0.22, 0.94) |
|  | β_e2_ | -0.19 (-0.38, 0.04) | -0.11 (-0.38, 0.15) | -0.40 (-0.57, -0.11) |

Note: path coefficients or β estimation without confidence intervals was due to infeasible non-linear constraint.

Supplemental Table 10: The relationship between comparison factors and the self-reported CES-D score using linear regression

| **Comparison variables (Buffer) ^b^** | **Coefficient (95% CI) ^a^** | | |
| --- | --- | --- | --- |
|  | Sex-combined | Male | Female |
| Mean tree cover  density (100 m) | -0.05 (-0.14, 0.05) | -0.07 (-0.20, 0.06) | -0.03 (-0.17, 0.11) |
| Percentage of impervious area (100 m) | 0.02 (-0.04, 0.09) | 0.11 (0.02, 0.21)* | -0.03 (-0.12, 0.06) |

* P<0.05

^a^ Adjusted for age, sex, education, work, marital status, smoking, alcohol drinking, illicit substance use, physical activity, and neighborhood deprivation score

^b^ 23 participants (9 males and 14 females) missing information on comparison variables

CI: confidence interval

Supplemental Table 11: The relationship between visibility factors and the self-reported CES-D score stratified by living in urban areas or not using linear regression

| **Visibility factor** | **Adjusted Coefficient (95% CI) ^a^** | | |
| --- | --- | --- | --- |
|  | Sex-combined | Male | Female |
| *Living in urban areas (individual twin n=476)* | | | |
| Sky | 0.15 (-0.17, 0.47) | -0.07 (-0.47, 0.34) | 0.36 (-0.12, 0.83) |
| Tree | -0.16 (-0.42, 0.10) | -0.15 (-0.55, 0.25) | -0.25 (-0.60, 0.11) |
| Building | 0.05 (-0.15, 0.26) | 0.15 (-0.16, 0.46) | 0.01 (-0.27, 0.28) |
| *Living in non-urban areas (individual twin n=1391)* | | | |
| Sky | 0.03 (-0.13, 0.19) | 0.06 (-0.17, 0.29) | 0.02 (-0.20, 0.25) |
| Tree | -0.01 (-0.15, 0.13) | -0.12 (-0.33, 0.08) | 0.06 (-0.14, 0.26) |
| Building | -0.07 (-0.41, 0.26) | 0.35 (-0.12, 0.82) | -0.33 (-0.80, 0.15) |

^a^ Adjusted for age, sex, education, work, marital status, smoking, alcohol drinking, illicit substance use, physical activity, and neighborhood deprivation score

CI: confidence interval
